# Racial-Ethnic and Gender Inequities in School SEND and Youth Justice Responses to Children and Young People’s Mental Health: Population-based Cohort Study using Linked Administrative Data in England

**DOI:** 10.64898/2026.07.13.26357949

**Authors:** Sorcha Ní Chobhthaigh, Twyla Greenway-Bailey, Josephine Musanu, Camille Cox, Mel Green, Amie Buhari, Cyra Neave, Mina Mawi, Aqsa Suleman, Rochelle A. Burgess, Matthew A. Jay, Delan Devakumar

## Abstract

**Background:** Mental health difficulties increase risk of justice system contacts, and justice involvement increases risk of and exacerbates existing mental health difficulties. However, discrimination influences both whether children’s needs are supported and the likelihood of justice system contacts. Examining patterning in Special Educational Needs and Disability (SEND) provision for Social, Emotional and Mental Health (SEMH) needs and youth justice contacts may identify escalation pathways and opportunities for equitable support.

**Methods:** In partnership with young people and community stakeholders, we analysed population-level linked school and police administrative data for 3.7m children (born 1997-2003) who attended state schools in England. We examined rates, timing and risk ratios for the sequencing of SEND-for-SEMH and youth justice contacts.

**Results:** Children ever recorded with SEND-for-SEMH had markedly elevated rates of youth justice contact, and vice versa. First justice contacts clustered between ages 14-16, though Black Caribbean, Mixed White-Black Caribbean, Romani, and Irish Traveller boys experienced steeper increases from age 12. Most racially and ethnically minoritised boys, alongside Romani and Irish Traveller girls, were significantly more likely than White British boys to have youth justice contacts before/without SEND-for-SEMH. Following SEND-for-SEMH, racially and ethnically minoritised boys and Irish Traveller girls were at greater risk of contacts.

**Conclusions:** Findings reveal intersectional discrimination in criminalisation of children and shed light on the complex bi-directional relationship between identified mental health need and youth justice contacts. A healing-centred, public health, multi-agency approach is urgently required to break the vicious cycle between unmet mental health need and youth justice system contacts.

## Introduction

Mental health difficulties, particularly externalising difficulties, increase risk of justice system contacts, and simultaneously, contacts with the justice system increase risk of mental health difficulties (1–4). Discrimination compounds this: increasing risk of mental health difficulties, influencing need identification and support provision through schools, access to and experiences of care, as well as policing practices and justice system processes (5–11). This cycle cumulates in escalating unmet need and perpetuating inequalities across health, education and justice systems.

Special Educational Needs and Disability (SEND) provision for Social, Emotional and Mental Health (SEMH) needs is the formalised channel for identifying ongoing emotional or behavioural difficulties in education (12). Intersectional discrimination exists in recorded SEND-for-SEMH, indicating both over-pathologisation of certain children and under-recognition of needs among others (7,13). Identification of need does not inherently result in appropriate support; the absence of which harms learning, perpetuates stigmatisation and worsens mental health (14–16). These unmet needs, failures and neglect by the school system, place racially and ethnically minoritised children at increased risk of youth justice contacts. These interactions with police are also detrimental to children’s mental health, sleep, academic performance and school engagement, reflecting potentially traumatic events for children and their families (1,2,17). However, as these contacts are neither random nor isolated, some children are more likely to be surveilled, experience discrimination in those interactions, and anticipate future negative interactions (5,18,19). Additionally, policing in schools, discrimination in school disciplinary practices and threats of police involvement mean these experiences extend into educational settings, with school becoming a potential source of harm (7,20–22).

As mental health inequalities continue to widen, and both SEND and Youth Justice systems undergo reform, understanding children’s experiences with policing, particularly children recognised as experiencing mental health need, is central to addressing avoidable harms. In partnership with young people and community stakeholders, we examine systematic differences in youth justice system contacts and cross-system pathways with SEND-for-SEMH based on gender and racial-ethnic group using population linked administrative data in England.

## Methods

### Data

We analysed linked de-identified administrative data from the Police National Computer (PNC) and National Pupil Database (NPD). The PNC contains information on recordable offences (cautions and convictions) in England and Wales. NPD contains information about pupils in state-maintained schools and other state-maintained educational settings in England.

PNC and NPD data were linked in 2019 as part of the Ministry of Justice (MoJ) and Administrative Data Research (ADR) UK funded data linkage program, Data First(23). Linkage was performed using a deterministic approach agreed between the MoJ and Department for Education (DfE). Data access was granted by the data owners, MoJ and DfE, February 2024. Study-specific ethical approval was obtained from UCL Research Ethics Committee (24643/001).

All data cleaning, preparation and analyses were completed with R software for Windows, version 4.4.0 (24) in the Office for National Statistics Secure Research Service (ONS SRS).

### Study Population

We included all (on-roll) pupils born 1997-2003 who attended state-school in England at any point during mandatory school years (age 5-16).

We defined a child’s NPD enrolment period from either the start of mandatory schooling (age 5) or the start of the school year of their first school census recording through to the end of mandatory schooling (age 16) or the end of the school year after their last census recording. Youth justice contacts started age 10 (the age of criminal responsibility in England) through to the child’s 18^th^ birthday.

### Exposure

Using an intersectional lens (25), we examined outcomes at the intersection of racialisation and gender, recognising the cumulative, rather than independent or additive, experiences. We operationalised racialisation using the NPD recording of “ethnicity” as a proxy for the lived experiences of individuals from ethnically and racially minoritised groups living in a White-British majority country. We use the terminology racial-ethnic groups given the Office for National Statistics Census categories reflect both “racial” and “ethnic” groups which are neither mutually exclusive nor does one inherently imply the other (26). Due to small numbers, we merged ‘Asian Other & Chinese’ groups but retained 16 other categories (Appendix Table 1). Where pupils had more than one value recorded, we used the most frequent. Pupils without “ethnicity” data (n=18,355) were excluded.

**Table 1.**
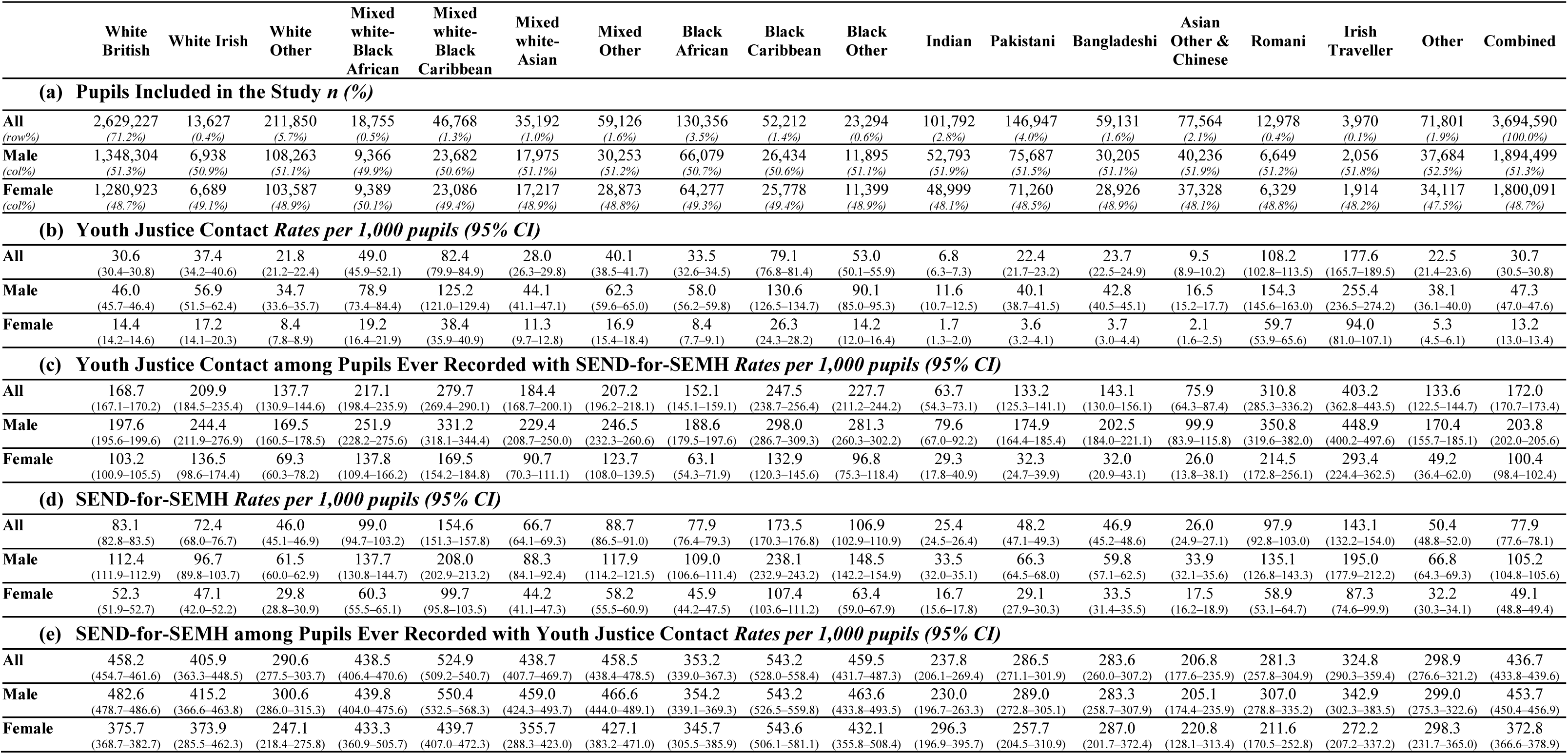
Cohort Overview & Outcome Rates per 1,000 pupils. (a) Pupils Included in the Study *n (%)* (b) Youth Justice Contact *Rates per 1,000 pupils (95% CI)* (c) Youth Justice Contact among Pupils Ever Recorded with SEND-for-SEMH *Rates per 1,000 pupils (95% CI)* (d) SEND-for-SEMH *Rates per 1,000 pupils (95% CI)* (e) SEND-for-SEMH among Pupils Ever Recorded with Youth Justice Contact *Rates per 1,000 pupils (95% CI)*

Gender was recorded in NPD as male or female, representing assigned “sex”, not gender identity. Where pupils had more than one value recorded, we used the most frequent.

### Outcomes

The two primary outcomes were NPD recording of SEND for mental health needs and youth justice contacts documented in PNC.

NPD SEND for mental health needs were indicated by records of SEND for Behaviour, Emotional & Social Difficulties (BESD) up to 2013/2014 and Social, Emotional and Mental Health (SEMH) difficulties from 2014/2015 onwards. BESD captured emotional and behavioural difficulties prior to the updated SEND code of practice (12,27) categorised under SEMH following this. We use SEND-for-SEMH to refer to any SEND for emotional and behavioural needs during the study time frame.

We use ‘youth justice contacts’ to refer to any documented PNC record of a caution or conviction for a recordable offence between age 10-17. This does not include interactions with police that did not result in a formal caution or conviction, such as ‘stop and search’ or the presence of police in schools.

### Statistical Analysis

#### Rates

We calculated rates per 1,000 pupils of ever being recorded with (a) youth justice contact among all pupils (b) youth justice contact among pupils ever recorded with SEND-for-SEMH (c) SEND-for-SEMH during their enrolment period among all pupils and (d) SEND-for-SEMH among pupils ever recorded with youth justice contact. Sensitivity analyses used enrolment person-time.

#### Timing

We examined timing trends using median age-at-first recording and time-to-event analyses using Kaplan-Meier curves to capture the cumulative incidence of first youth justice contact each year from age 10 and SEND-for-SEMH from enrolment. Age was calculated using an approximate date of birth (15^th^ of birth month) derived from NPD.

#### Pathways

We calculated risk ratios for the likelihood that a pupil would be recorded with youth justice contact first, versus either no such record or a recording of SEND-for-SEMH first. We repeated analyses to ascertain the likelihood of SEND-for-SEMH first. For those identified with SEND-for-SEMH first, we calculated the risk ratios of a subsequent youth justice contact. For boys first recorded with youth justice contact, we calculated the risk ratios of subsequently being recorded with SEND-for-SEMH. To supplement these analyses, where possible, we conducted logistic regression modelling with an interaction term to examine compounded impacts at the intersection of racial-ethnic group and gender. Instances where the order of recording patterns could not be determined were removed, i.e. youth justice contacts dated within the 4-months period between two census recordings (n=1,754; 71.8% White British, consistent with cohort composition).

We used White British males as the reference group for theoretical and practical reasons. Theoretically, without equity-oriented policies, the White British majority experience by default becomes the normative expectation against which minoritised groups are compared (28). White British males represent the largest group in SEND-for-SEMH (29,30) and policing data (31). This approach allows us to quantify how the experiences of others differ from this implicit ‘norm’, rather than suggesting White British males represent an ideal standard.

The effects of socioeconomic position are intertwined with historical and ongoing experiences of discrimination, oppression and exclusion, including, based on race, ethnicity or migration status, which contribute to lower socioeconomic positioning for minoritised groups and collectively impact health, education and justice outcomes (32). Adjusting for socioeconomic position risks diluting the impact of systemic inequalities on the outcomes of interest, therefore, we report unadjusted results only.

### Participation

Young people with lived experience of racialisation and navigating mental health difficulties during their school years, and community stakeholders, guided protocol development and shaped the interpretation of results. Young people engaged in shared decision-making throughout analysis and study refinement. Collaborative discussions led to the development of co-produced recommendations.

## Results

### Study cohort

Our cohort included 3,694,590 pupils born 1997-2003 who attended state-maintained schools in England between 5-16 years (Table 1). See Supplementary Figure 1 for data flow summary.

### Rates

Rates of youth justice contacts per 1,000 varied significantly by gender and racial-ethnic group (Figure 1a; Table 1). Within each racial-ethnic group, boys had higher rates than girls, though rates for some girls exceeded those of boys from other groups. Rates were highest for pupils from Irish Traveller, Romani, Black Caribbean, and Mixed White-Black Caribbean backgrounds while those from Indian and Asian Other & Chinese groups had the lowest rates. Trends remained stable in sensitivity analyses (Supplementary Table 3).

**Figure 1:**
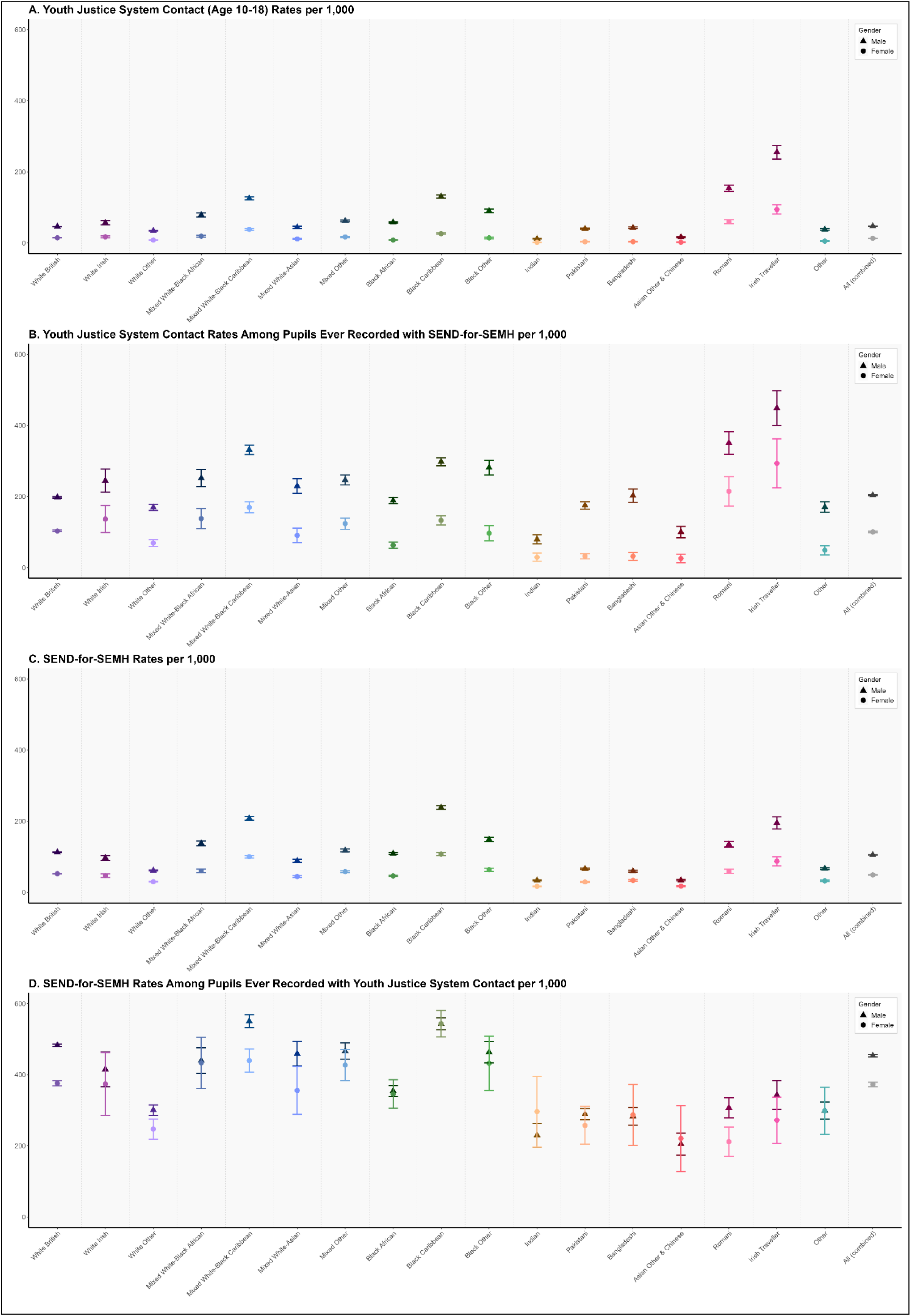
Rates per 1,000 pupils of ever being recorded with (a) Youth Justice Contact (b) Youth Justice Contact among Pupils Ever Recorded with SEND-for-SEMH (c) SEND-for-SEMH (d) SEND-for-SEMH among Pupils Ever Recorded with Youth Justice Contact

Rates of SEND-for-SEMH showed substantial variation by racial-ethnic group and gender (Figure 1c; Table 1). Similarly, within racial-ethnic groups boys had higher rates than girls, though the magnitude of this gap varied. Rates were highest for Black Caribbean, Mixed White-Black Caribbean and Irish Traveller pupils, and lowest among those from Asian, White Other, and Other racial-ethnic backgrounds. Trends remained stable in sensitivity analyses, with higher rates among Romani pupils when accounting for school enrolment person-time.

Rates of youth justice contacts were markedly higher among pupils ever recorded with SEND-for-SEMH, and rates of SEND-for-SEMH were correspondingly higher among pupils ever recorded with youth justice contacts, reflecting the same pattern of co-occurrence from both systems’ vantage points (Figures 1b & 1d; Table 1). Rates among girls were 7.6x higher than the cohort baseline (range: 3.1x – 17.7x) and among boys 4.3x higher (range: 1.8x – 6.9x), with the largest increases observed among Indian and Asian Other & Chinese pupils.

### Timing

Although median age at first youth justice contact across genders clustered from age 14 to 16, trends differed for girls and boys (Figure 2; Supplementary Tables 4 & 5). Girls had higher incidences of first contacts between ages 14-16, with less pronounced increases in later teenage years. While the curves for girls from Asian groups were relatively flat, increases were more pronounced for Mixed White-Black Caribbean and Black Caribbean girls, and most pronounced for Irish Traveller and Romani girls who experienced younger and more consistent increases across ages.

**Figure 2:**
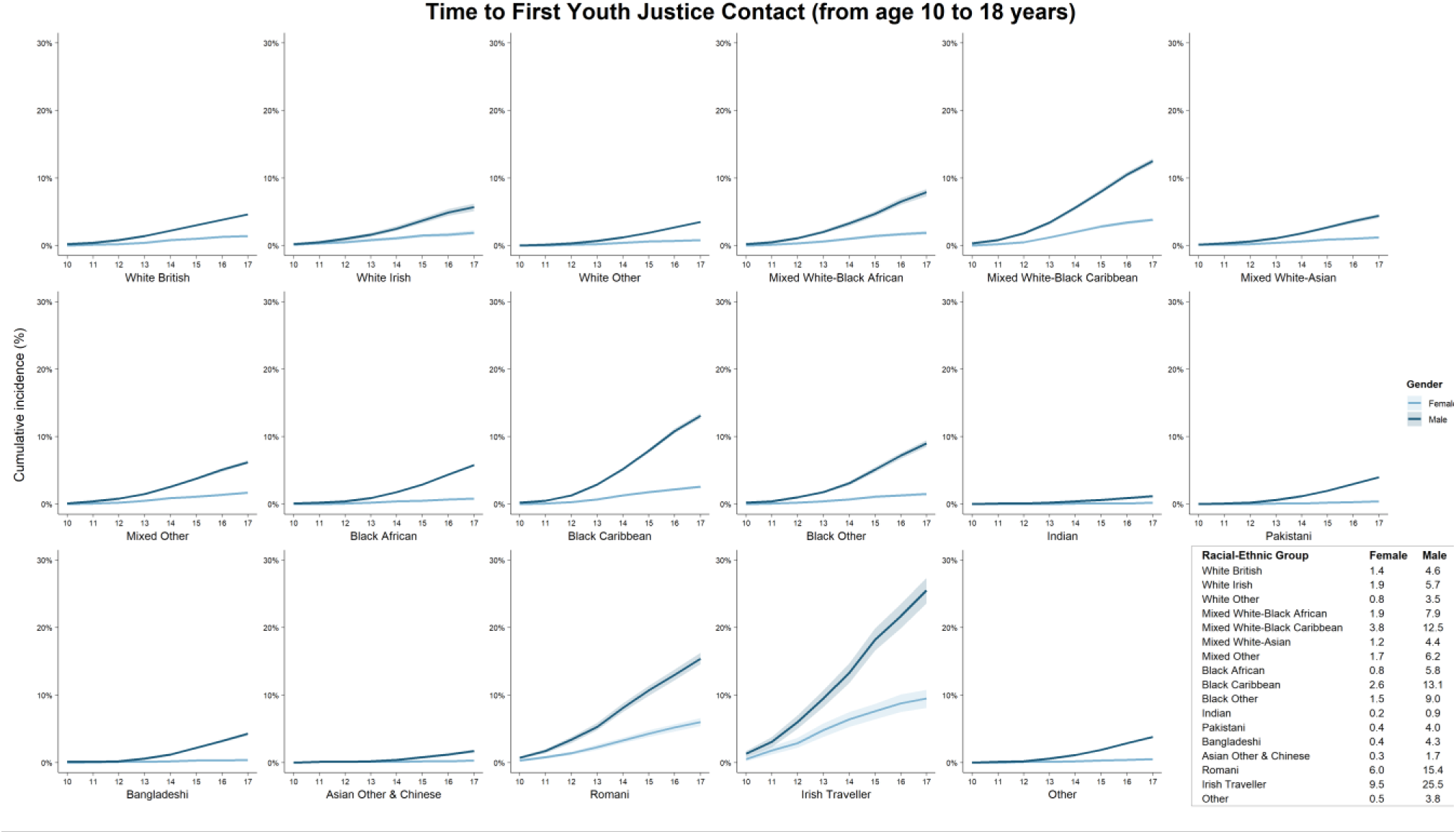
Time-to-first Youth Justice Contact Kaplan-Meier Curves. Curves illustrate the proportion of pupils with a first recording each year, with steeper rises indicating higher recordings. Inset table shows cumulative incidence (%) at end of 17^th^ year by racial-ethnic group and gender.

Among boys, timing trends varied markedly across racial-ethnic groups. Black Caribbean, Mixed White-Black Caribbean, Romani and Irish Traveller boys experienced steep increases from age 12, most pronounced between ages 14-16. A similar, although less pronounced trend was found for Mixed White-Black African and Black Other boys. Curves for White and Mixed White-Asian boys show a steady increase across adolescence, those for Black African, Pakistani, Bangladeshi and Other racial-ethnic boys appear relatively flat until age 14, while increases for Indian and Asian Other & Chinese boys remain low.

SEND-for-SEMH age and time trends varied considerably across groups (Supplementary Figure 2; Tables 4 & 6). Across racial-ethnic groups, boys were recorded younger and more steadily from enrolment than girls. Boys from Irish Traveller, Black Caribbean, Mixed White-Black Caribbean and Black Other backgrounds experienced a higher incidence in early years. Mixed White-Black Caribbean, Black Caribbean, Romani and Irish Traveller girls also experienced a higher incidence and steadier increase relative to other girls, while curves for most girls indicated a slow and gradual increase until later school years. Curves for boys and girls from Indian and Asian Other & Chinese groups were relatively flat, reflecting very slight yearly increases through the enrolment period.

### Pathways

Compared to White British boys, boys from White Irish, Mixed White-Black, Mixed Other, Black, Pakistani, Bangladeshi, Romani, Irish Traveller and Other unspecified racial-ethnic backgrounds were significantly more likely to have youth justice contacts first relative to SEND-for-SEMH first or no record in either system (Figure 3a). While White Other and Mixed White-Asian boys were not significantly different, Indian and Asian Other & Chinese boys were less likely. Nearly all girls were significantly less likely to be recorded with contacts first, except for Romani and Irish Traveller girls who were significantly more likely.

**Figure 3:**
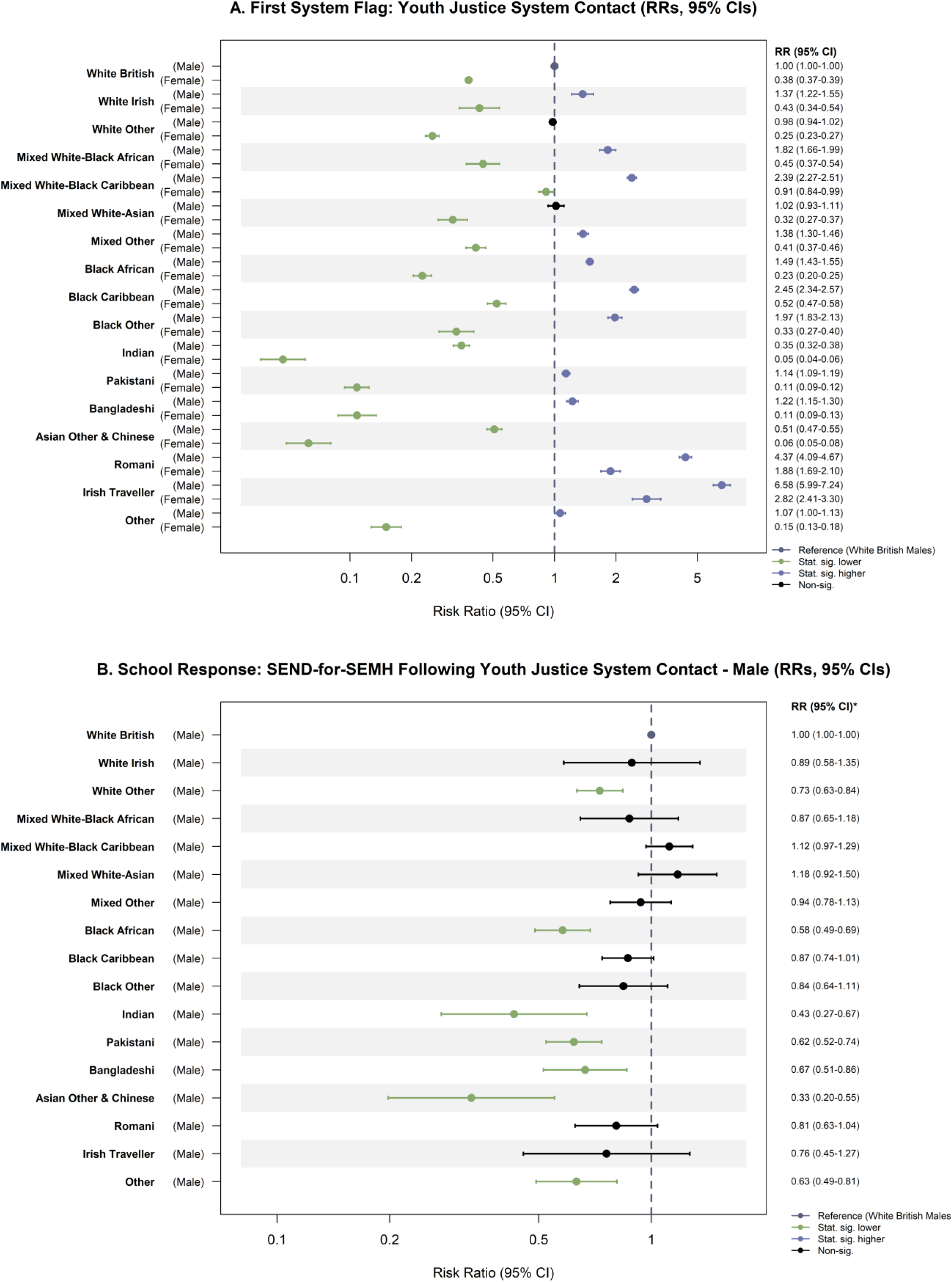
Youth Justice Contact Pathway. Relative risk, by racial-ethnic group and gender of (a) youth justice contact being the first recorded indicator of need (vs. SEND-for-SEMH first or neither) (b) among boys with youth justice contact first, a subsequent recording of SEND-for-SEMH

Of boys recorded with youth justice contacts first during their enrolment period, those from Black African, Asian, White Other and Other racial-ethnic groups were significantly less likely to subsequently receive SEND-for-SEMH (Figure 3b).

Regression interaction testing confirmed significant intersectional effects of racial-ethnic group and gender on likelihood of youth justice contact first (p<0.0001) (Supplementary Table 7). For youth justice-to-SEND-for-SEMH, censoring restricts reporting of the interaction testing and main effects by racial-ethnic group.

Compared to White British boys, Mixed White-Black, Mixed Other, Black Caribbean, Black Other, Romani and Irish Traveller boys were significantly more likely to be identified with SEND-for-SEMH first relative to no record in either system or a youth justice contact first (Figure 4a). All other boys and girls were significantly less likely to be recorded with SEND-for-SEMH first.

**Figure 4:**
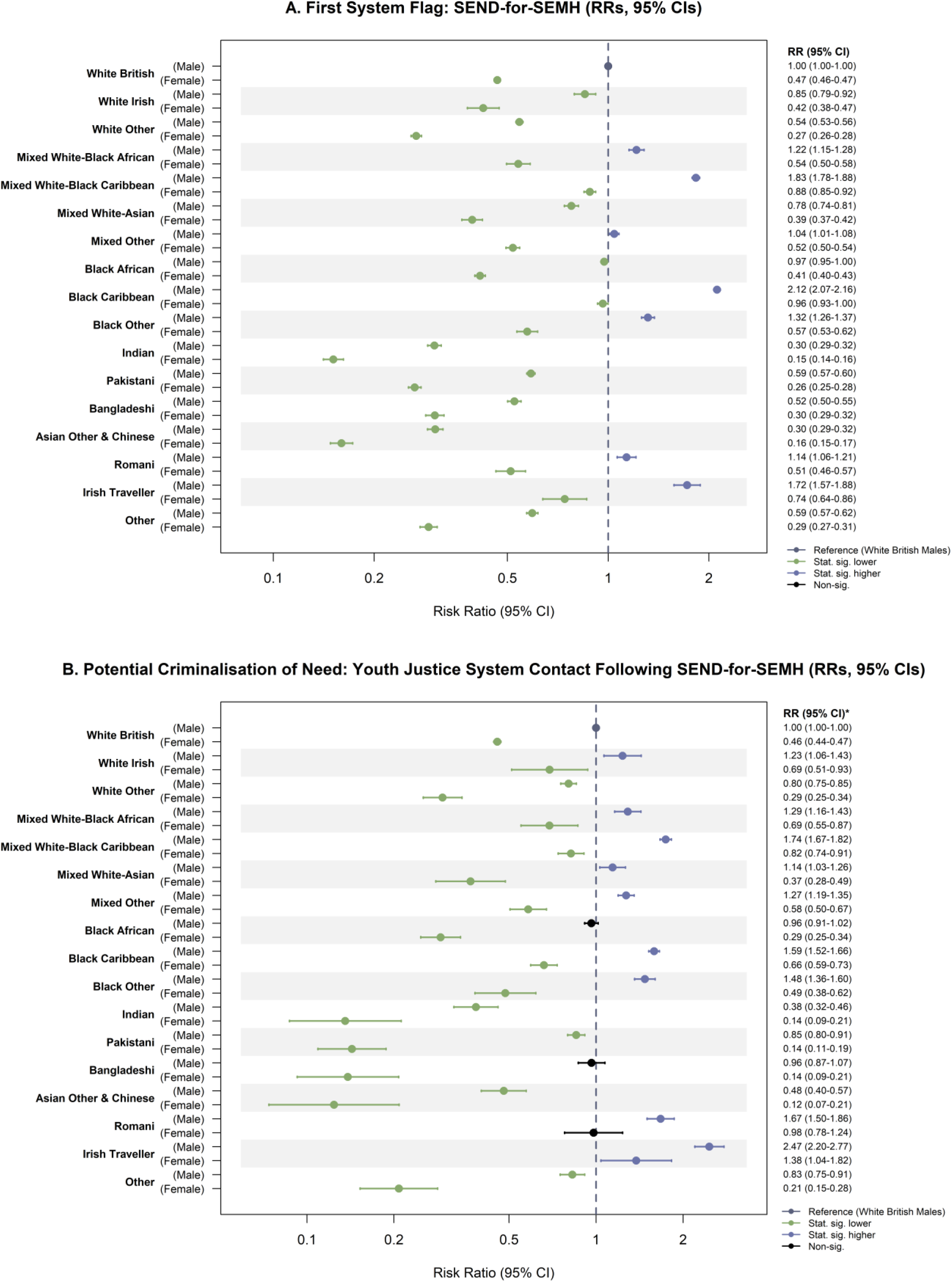
School SEND-for-SEMH Pathway. Relative risk, by racial-ethnic group and gender of (a) SEND-for-SEMH being the first recorded indicator of need (vs. youth justice contact first or neither) (b) among boys with SEND-for-SEMH first, a subsequent youth justice contact.

Of those recorded with SEND-for-SEMH first, boys from White Irish, Mixed racial-ethnic, Black Caribbean, Black Other, Romani and Irish Traveller backgrounds were significantly more likely to have a subsequent youth justice contact (Figure 4b). Nearly all girls were significantly less likely to have subsequent youth justice contact, except for Irish Traveller girls who were significantly more likely and Romani girls who were no more or less likely than White British boys.

Regression interaction testing confirmed significant intersectional effects of racial-ethnic group and gender on likelihood of being recorded with SEND-for-SEMH first (p<0.0001) and of subsequent youth justice contact (p<0.0001) (Supplementary Table 7).

## Discussion

This study provides the first comprehensive examination of intersectional patterns and pathways by racial-ethnic group and gender in youth justice contacts and mental health-related SEND provision using population-level linked administrative data. Rates of youth justice contacts were substantially higher among boys and girls from Black Caribbean, Mixed White-Black Caribbean, Romani and Irish Traveller backgrounds alongside Mixed White-Black African and Black Other boys, with higher rates among these same groups in SEND-for-SEMH. Rates of youth justice contacts were higher among pupils ever recorded with SEND-for-SEMH and vice versa, reflecting the co-occurrence of formally documented mental health need and justice system involvement before turning 18. This pattern was most pronounced among girls and Asian, Black African, White Other and Other racial-ethnic groups who have lower baselines and are consistently least likely to receive formalised school provision despite mental health-related hospital contacts or need indicators (7,13), suggesting heightened vulnerability among those identified.

Our findings highlight the ‘critical period’ during the early teens, with documented impact across health and education outcomes following the transition to secondary school (33). First youth justice contacts cluster between ages 14-16, however, Black Caribbean, Mixed White-Black Caribbean, Romani, and Irish Traveller boys and girls appear criminalised younger, with marked escalation from age 12. Girls show more concentrated risk between 14-16 years, indicating a critical period of vulnerability. Boys from White Irish, Mixed White-Black, Mixed Other, Black, Pakistani, Bangladeshi, Romani and Irish Traveller backgrounds – and girls from Romani and Irish Traveller backgrounds - are significantly more likely than White British boys to encounter youth justice system first, either before or without SEND-for-SEMH. Boys from Asian, Black African, White Other and Other unspecified racial-ethnic groups are significantly less likely to be recorded with SEND-for-SEMH following youth justice contacts. Even following formal identification of mental health need, boys from White Irish, Mixed racial-ethnic, Black Caribbean, Black Other, Romani backgrounds and both Irish Traveller boys and girls remain at elevated risk of criminalisation.

Mental health impacts learning, academic performance, relationships and risk of youth justice involvement (4,34). Inversely, police and justice system contact increases risk of mental health difficulties, with impacts to education engagement, academics, and relationships with public systems (1–3,17,18). Our findings evidence this complex bi-directional relationship, however, risk patterning systematically differs at the intersection of racialisation and gender, reaffirming systematic differences in how different presentations, emotional or behavioural needs, are interpreted and responded to across systems.

For girls, vulnerability appears particularly salient from age 13, aligning with timing of mental health-related hospital contacts (29). While girls are less likely to receive formalised school provision after indicators of mental health need and hospital contacts (7,13), our findings indicate those who are identified with SEND-for-SEMH are at heightened risk of youth justice contacts, suggesting they may be experiencing significant distress or vulnerability. Girls’ needs are often compounded and neglected by interactions with the justice system built on rigid gendered stereotypes and punitive responses to perceived ‘deviance’ (35,36). Mixed White-Black Caribbean, Black Caribbean, Romani and Irish Traveller girls experience contacts younger and more steadily than their peers – comparative to or even higher than some of the male groups. This aligns with differing custodial placements experienced by racialised girls, likely reflecting biased perceptions of need compounded by adultification (35,37).

While girls and specific racial-ethnic groups face under-recognition of need, other racially and ethnically minoritised children, particularly boys, simultaneously experience over-policing and ongoing unmet need. Consistent with documented discriminatory policing practices (11,38), racially and ethnically minoritised children are more likely to have youth justice contacts first and from a younger age, suggesting these interactions may precede, precipitate, or perpetuate need. In particular, Black Caribbean, Mixed White-Black Caribbean, Romani, and Irish Traveller children appear criminalised from the age of 12, aligning with reports of over-pathologisation and over-surveillance (20,39). Racialised stereotypes, such as perceptions of Black children as aggressive or Irish Travellers as criminal, alongside adultification biases, likely shape how these children’s presentations are interpreted and responded to (40,41).

Inequities in need identification and youth justice contacts reinforce barriers to accessing appropriate care. Racially and ethnically minoritised parents’ report feeling dismissed, judged, and excluded from decision-making when seeking support from schools (20,42) further highlighting missed opportunities for restorative early interventions. When obtained, formal identification of mental health needs may flag vulnerability but fails to translate into appropriate support. Community policing strategies that increase police presence in communities and schools with higher deprivation and ethnic density (22) converge with inequities in school responses to need and disciplinary practices (7). This convergence of discriminatory school and policing practices likely fuels a cycle of escalating need, hypervigilance, distrust in and disengagement from public services, deterring help-seeking - especially when vulnerability is recognised but not addressed. This risks reinforcing deficit narratives and racist beliefs, undermining child self-esteem and aspirations, funnelling children into the school-to-prison pipeline, rather than understanding children’s needs and implementing effective, healing-centred, restorative, culturally sensitive interventions.

Under the Equality and Education Acts, schools are required to protect pupils from discrimination and ensure school environments are safe - physically, emotionally and culturally (43,44). Meeting these duties requires equitable responses, however, national policy currently lacks coherent guidance on mental health, cultural safety and restorative approaches (45) to adequately equip school providers to perform these duties and ongoing failures to explicitly address inequalities in policy perpetuate inequitable practices.

Strengths include the novel use of longitudinal national linked administrative data from state school and police records to examine equity in system responses to children and pathways between formally identified mental health needs and youth justice contacts in England. Completed in partnership with young people and community stakeholders, this work integrates meaningful participation in quantitative analysis. Sensitivity analyses using person-time confirmed trends. Examination of regional and school-level variation was outside the scope of this analysis, though are likely influenced by overlapping socio-political-economic factors. While reporting more granular racial-ethnic groups than previous research, aggregated categories inevitably invisibilise some experiences and groups; all “Other” categories mask diverse experiences, some communities (e.g. Latinx, Arab) lack formal recognition and many individuals’ identities do not fit prescribed categories. Our findings are likely a conservative underestimate as PNC does not capture informal interactions that did not result in a formal caution or conviction - such as ‘stop and search’ or police officers involved in school discipline - with documented inequities in these practices (22,46). Additional research is needed to explore of the impacts of informal contacts on mental health, relationships with school and help-seeking. Understanding parental experiences of discrimination in education and the influence on school-family relationships, children’s educational experiences and longer-term outcomes is critical to informing intergenerational approaches to healing from public system harms. Further examination of school-based indicators of need in relation to youth justice will provide insight into the escalating cycle and opportunities for equity-oriented responses to shift trajectories of risk.

In the context of well-documented racist policing practices (11), our results add to the qualitative reports (20)that pathways to criminalisation reflect systematic bias in how children’s needs are interpreted and responded to, rather than individual children, specific providers or isolated practices; some children are policed as adults, while others are allowed to experience adolescence. The convergence of discriminatory practices across systems puts children ‘at risk’ directly through harmful practices and indirectly by failing to appropriately identify and support need. While all children are negatively impacted by these failures, the cumulative burden of harm falls most heavily on Black, Mixed White-Black, Romani and Irish Traveller children, historically ostracised communities, whose distress continues to be mislabelled as deviance and who cannot wait any longer for incremental reform. Crucially, addressing these failures requires a public health multi-system approach, co-ordinated across education, health, social care and justice systems - an effective strategy cannot treat these separately.

Creating mentally healthy systems demands those working with children are adequately supported to have the knowledge, skills and capacity to do so; this requires addressing gaps in current policy, guidance, and co-ordination. Coherent evidence-based policy directives are needed to resolve current contradictions in how children’s behaviour is understood and responded to across systems, whether as an indicator of unmet need or as grounds for sanction. Clear guidance must be established for all public sector providers interacting with children under 18 covering mental health to build shared foundational understanding, contextual safeguarding to address environmental triggers, and evidence-based protocols to enable safe, restorative and healing-centred responses. This includes equipping police to interact safely with children, minimising risks of harm and (re)traumatisation. Equity-oriented policy that embeds training and accountability into implementation is critical to rectifying systemic biases and enabling individuals within these systems to perform their duties safely, effectively and equitably. Anti-discrimination and cultural safety training, including addressing implicit bias, racialised and gendered stereotypes, must be integrated into compulsory training and qualification requirements for all providers. Accountability mechanisms are central to just and fair public systems and to identifying gaps; ongoing inequities monitoring, Written Statements of Action, and transparent complaints processes are all crucial to sustaining improvements. To address co-ordination failures, a formalised cross-system ‘care pathway’ is needed, with key worker oversight to ensure equitable access to assessment, support and diversion programs. Investment in and partnerships with community organisations will be integral, both to providing prevention and early intervention that reduces reliance on statutory services, and to building trust and acting as intermediaries for communities consistently failed by the state across generations.

A public health approach must be recognised as essential, not aspirational, to fulfil legal requirements under the Public Sector Equality Duty and Safeguarding Duty of Care, to protect children from harm and to ensure they can access their rights to education and health.

## Ethics

This study uses linked, de-identified data from the National Public Database and Police National Computer as part of the Ministry of Justice (MoJ) and Administrative Data Research (ADR) UK funded data linkage program, Data First. Data access was granted by the data owners, MoJ and DfE, February 2024. Study-specific ethical approval was obtained (10.03.2023) from UCL Research Ethics Committee (24643/001).

## Data availability

This study uses routinely collected administrative data from the Ministry of Justice (MoJ) and Department for Education (DfE). The data is not publicly available and can be accessed by researchers by applying directly to the MoJ and DfE.

## Author contributions

SNC had access to the data in the study and takes responsibility for the integrity of the data and the accuracy of the data analysis. SNC conceptualised and designed the study, performed the data analyses, and wrote the initial draft manuscript. CC, JM and TGB contributed to study design, analysis decisions, interpretation of results and co-development of recommendations. CN, MG, MM, AB and AS contributed to interpretation of results and co-development of recommendations. MJ supported data management, code development and supervised the analytical approach. RB supervised the participatory methodology. DD provided supervisory support throughout. All authors reviewed, commented, and approved the final manuscript.

## Acknowledgements

We would like to express our appreciation to additional and former stakeholder advisors (in addition to those who are named authors). Their meaningful contributions and thoughtful insights shaped this project, contextualisation and co-developed recommendations. We also extend our wholehearted gratitude to the No More Exclusions coalition for their valuable feedback and insights.

We gratefully acknowledge all children, young people and families whose de-identified data are used in this research.

This work was undertaken in the Office for National Statistics Secure Research Service using Data First linked administrative data. The use of the data in this work does not imply the endorsement of the ONS or data owners (e.g. MoJ and DfE) in relation to the interpretation or analyses of the statistical data. This work uses research datasets which may not exactly reproduce National Statistics aggregates. National statistics follow consistent statistical conventions over time and cannot be compared to Data First linked datasets.

The views in this publication do not necessarily reflect the views of UCL.

